# Differentiating between infectious and non-infectious influenza A virus and coronavirus RNA levels using long-range RT-qPCR

**DOI:** 10.1101/2021.11.11.21266219

**Authors:** Dovile Juozapaite, Charlotte V. Rigby, Ingrida Olendraite, Pankaj Mathur, Kalyan Dhanorkar, Vishalraj Hulle, Tejas Shah, Vijeta Jadhao, Shravan Mutha, Hamid Jalal, Vikram Gopal, Aartjan J.W. te Velthuis

**Affiliations:** Vilnius University Hospital Santaros Klinikos, Hematology, Oncology and Transfusion Medicine Center, Santariskiu st. 2, 08661 Vilnius, Lithuania; Lewis Thomas Laboratory, Department of Molecular Biology, University of Princeton, Princeton, 08540 NJ, United States; University of Cambridge, Department of Pathology, Addenbrooke’s Hospital, Cambridge CB2 2QQ, United Kingdom; Public Health England, Addenbrooke’s Hospital, Cambridge CB2 2QQ, United Kingdom; Pune Instrumentation Pvt. Ltd., PCNTDA, Bhosari, Pune, 411026, India; Krsnaa Diagnostics Ltd, Chinchwad, Pune, 411033, India

**Keywords:** SARS-CoV-2, influenza A virus, RT-qPCR, UVC inactivation, long-range

## Abstract

During the Coronavirus Disease 2019 (COVID-19) pandemic, residual SARS-CoV-2 genome and subgenomic RNA fragments were observed in recovered COVID-19 patients. The presence of such RNAs in the absence of live virus leads to incorrectly positive RT-qPCR results, potentially delaying medical procedures and quarantine release. We here propose a simple modification to turn commercial COVID-19 RT-qPCR protocols into long-range RT-qPCR assays that can differentiate between infectious and non-infectious influenza and coronavirus RNA levels. We find that the long-range RT-qPCR method has a sensitivity that is indistinguishable from a commercial Taq-Path COVID-19 RT-qPCR assay when tested on clinical samples taken withing 5 days of the onset of symptoms. In clinical samples taken at least 15 days after the onset of symptoms when patients had recovered from COVID-19, the modified RT-qPCR protocol leads to significantly fewer positive diagnoses. These findings suggest that the long-range RT-qPCR method may improve test-to-release protocols and expand the tools available for clinical COVID-19 diagnosis.

**Importance:** Various molecular tests can detect viral RNA in clinical samples. However, these molecular tests cannot differentiate between RNA from infectious viruses or residual viral genome fragments that are not infectious. In several percent of COVID-19 patients, such residual viral RNAs can be detected long after recovery and the disappearance of infectious SARS-CoV-2. These “persistently-positive” RT-qPCR results are different from false-positive RT-qPCR results, which can be generated due to in vitro cross-reactivity or contaminations. However, the detection of RNA fragments leads to incorrect conclusions about the status of a COVID-19 patient and an incorrect diagnosis. We here modified the commercial Taq-Path COVID-19 RT-qPCR kit to make this test less sensitive to residual viral RNA genome fragments, reducing the likelihood that incorrect RT-qPCR results affect the treatment or quarantine status of recovered COVID-19 patients.

## Introduction

Respiratory RNA viruses, such as influenza and coronaviruses, are important human pathogens that have a substantial impact on our healthcare systems and economy (1, 2). The detection of viral RNA infections in symptomatic and asymptomatic patients is essential for preventing respiratory virus spread and monitor patient recovery. Various assays exist to detect viral nucleic acids or proteins in clinical samples. Among these tests, reverse transcription quantitative polymerase chain reaction (RT-qPCR)-based methods are considered the gold-standard.

RT-qPCR assays consist of two steps: cDNA synthesis and PCR amplification (Fig. 1). In commercial kits, random hexamers prime the cDNA synthesis reaction. These primers provide flexibility and hybridize to the viral genome as well other RNA molecules present in the clinical sample, including host cell ribosomal RNAs, host cell and viral messenger RNAs, replicative intermediates of viral replication, and fragments of host cell and viral RNAs released by dead cells. To generate a specific signal from this complex pool of cDNA molecules, PCR primers next selectively hybridize to a cDNA copy of the viral genome to prime the amplification of a 50-250 nt long section of the cDNA by a heat-stabile DNA polymerase (Fig. 1).

**Figure 1.**
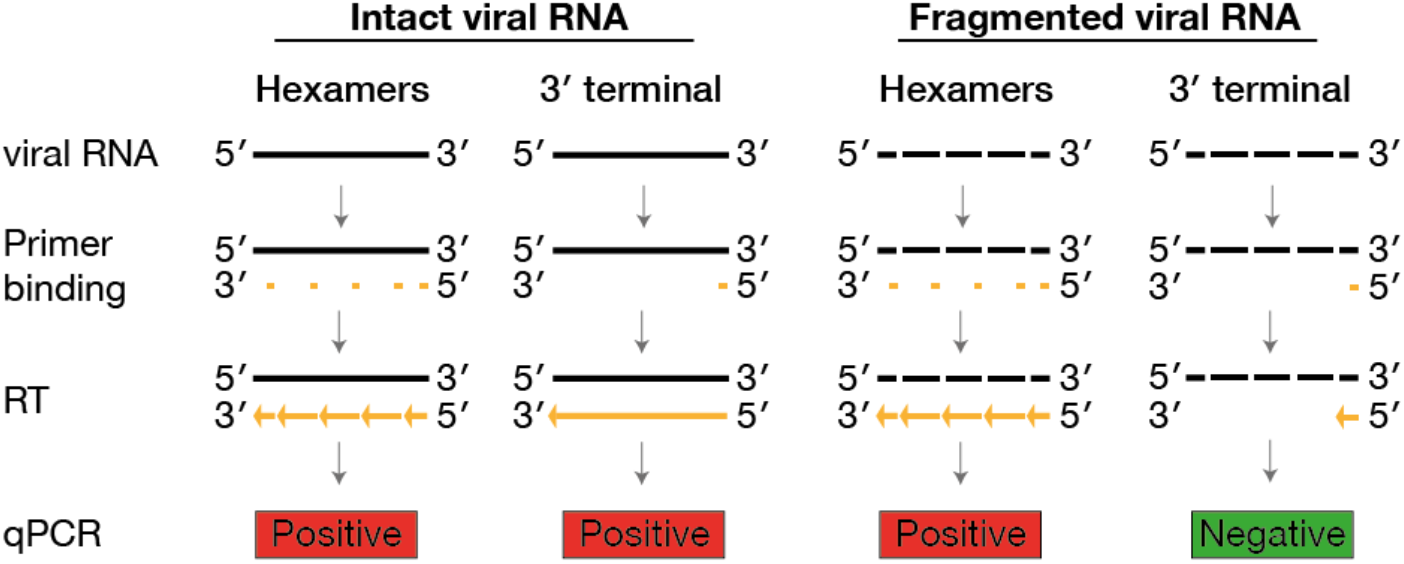
Schematic of differential RT-qPCR detection of intact or fragmented viral RNA using an RT step that is dependent on random hexamers or a primer binding to the 3′ terminus of the viral RNA.

The above procedure yields a signal that typically has excellent sensitivity and accuracy. However, many studies have reported that samples obtained from recovered Coronavirus disease 2019 (COVID-19) patients may still yield positive RT-qPCR results, even though no infectious virus can be cultured from these samples (3–5). These findings suggests that these RT-qPCR assays fail to distinguish between infectious and non-infectious viral RNA molecules. This hypothesis is supported by a recent human challenge study, which showed that lateral flow assays provide a better estimate of the infectious viral load than a commercial RT-qPCR assay (6). Since lateral flow assays are not high-throughput, alternative assays are needed to distinguish between infectious and non-infectious viral RNA levels in clinical samples.

Persistently positive RT-qPCR signals are likely derived from remnants of viral RNA that were converted to cDNA by random hexamers and the RT enzyme. To minimize the generation of cDNA molecules from such viral RNA remnants, we here replaced the random hexamers with an oligonucleotide that specifically binds to the 3′ end of the viral genome, thereby converting only intact viral genomes to cDNA products that can serve as qPCR template. Breaks in the viral genome would lead to early termination of cDNA synthesis and no cDNA amplification during the PCR stage (Fig. 1). We find that the long-range RT-qPCR method facilitates the differential detection of infectious and non-infectious influenza A and human coronavirus (HCoV) 229E, in agreement with previous influenza A virus studies (7, 8). Moreover, the RT-qPCR provides the same sensitivity as commercially available Taq-Path RT-qPCR kits at detecting severe acute respiratory syndrome coronavirus 2 (SARS-CoV-2) RNA in clinical samples, but the assay yields significantly fewer positive results in clinical samples obtained from clinically recovered COVID-19 patients. These findings suggest that the long-range RT-qPCR method can be used to exclude non-infectious from persistently positive COVID-19 cases.

## Results

### Detection of infectious and non-infectious influenza A virus

Ultraviolet C (UVC) can create cross-links or breaks in nucleic acid strands and inactivate influenza A virus (7, 9, 10), HCoV 229E, and SARS-CoV-2 (11–17). Following calibration of our UVC instrument (Fig. 2A), we exposed 25 µl 10^6^ pfu/ml influenza A virus strain A/WSN/1933 (H1N1; Genbank LC333182) to 11-54 W of UVC for 0 to 120 seconds (Fig. 2A). The exposed virus was subsequently serially diluted to determine the virus titer by plaque assay. We found that the virus sample was completely inactivated at 54 W UVC after 60 seconds of exposure (Fig. 2A). Viral RNA was next extracted from the 54 W time course samples, and cDNA generated using random hexamers or a universal influenza A virus primer (TUMI 12G) designed to bind to the conserved 3′ end of the viral genome segments (18, 19). cDNA levels were subsequently analyzed using primer sets specific for the influenza A virus M or NA segment (Fig. 2B and 2C, respectively). qPCR signals obtained with the 3′ end-derived cDNA showed an increase in Ct value that was strongly correlated with the virus titer and length of UVC exposure (Fig. 2C). No effect of UVC was observed on the viral nucleoprotein, which forms ribonucleoprotein complexes with the viral RNA (20) (Fig. 2B). In contrast, the qPCR signal obtained with random hexamers showed a poor correlation with the viral titer (Fig. 2C). These results suggest that a long-range RT-qPCR protocol provides the best estimate of the infectious influenza A virus titer than a protocol based on random hexamers, in line with previous influenza A virus studies (7, 8).

**Figure 2.**
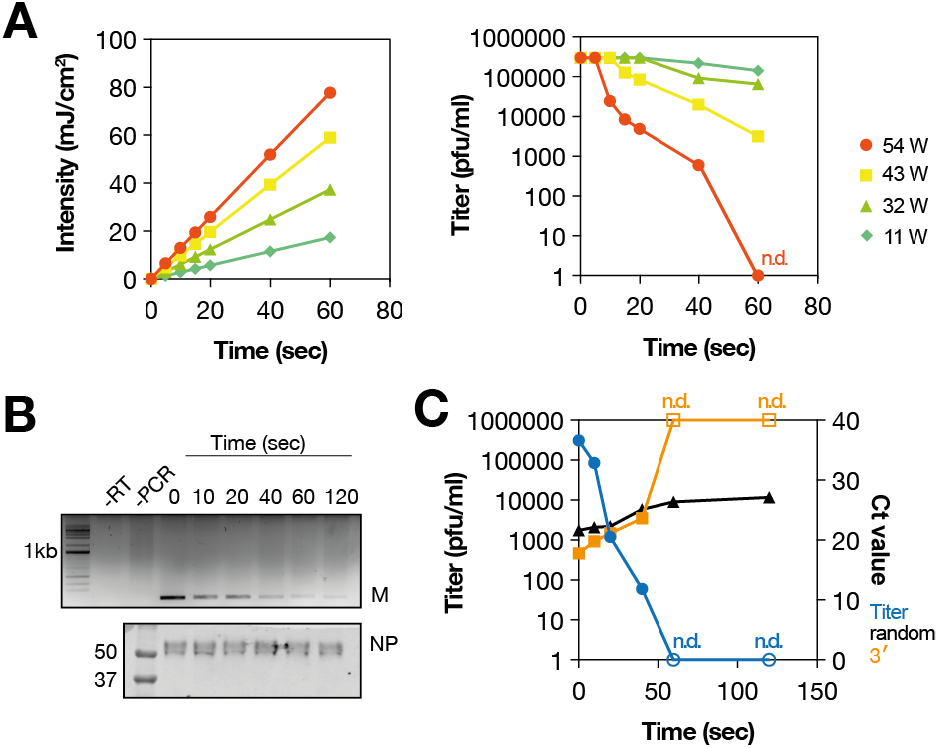
Detection of infectious and non-infectious influenza A virus RNA. (**A**) The UVC intensity was measured for every exposure (left) and the effect of UVC on the virus titer determined by plaque assay (right). Influenza A/WSN/1933 (H1N1) virus was placed on a plastic surface and exposed for 5 to 60 seconds at 11 to 54 W UVC. (**B**) Top - gel electrophoresis analysis of M segment RT-PCR signal after UVC exposure at 54W. The RT step was performed using a universal influenza A virus 3′ terminal RT primer. Bottom - western analysis using an antibody specific for the influenza A virus NP protein. (**C**) Relation between UVC exposure at 54 W, the influenza A virus A/WSN/1933 titer, and the qPCR signal following RT reactions with random hexamers or a universal influenza A virus primer capable of binding to the 3′ terminus of each of the eight viral RNA segments. qPCR was performed using primers specific for the NA segment. Plaque assay results that were not detectable (n.d.; open squares) are indicated. RT-qPCR signals that were not detectable are shown as Ct = 40 on graph.

### Detection of infectious and non-infectious human coronavirus 229E

To investigate whether the long-range RT-qPCR protocol can distinguish between infectious and non-infectious coronavirus samples, we exposed 25 µl 10^4^ pfu/ml HCoV 229E (Genbank AF304460) to 54 W UCV for 0 to 120 seconds (Fig. 3A). Determination of the virus titer by plaque assay showed a clear reduction in virus titer over the time-course. No viable virus was found after 60 seconds of exposure. Next, viral RNA was extracted, and cDNA synthesis performed using random hexamers or an oligo-dT_20_ primer that binds to the 3′ polyA-tail of the HCoV 229E genome. qPCR analysis on the two cDNA sample sets using primers targeting the N gene, as described previously (21), showed a strong correlation between the oligo-dT_20_ qPCR Ct values and the viral titer (Fig. 3A). In contrast, reactions containing cDNA generated using random hexamers showed a poor correlation between the qPCR Ct values and the viral titer (Fig. 3A). Together the above results suggest that a long-range RT-qPCR protocol that uses a primer that binds to the 3′ end of the coronavirus genome during the cDNA synthesis provides a better estimate of the infectious viral titer than RT-qPCR protocols that rely on random hexamers for cDNA synthesis.

**Figure 3.**
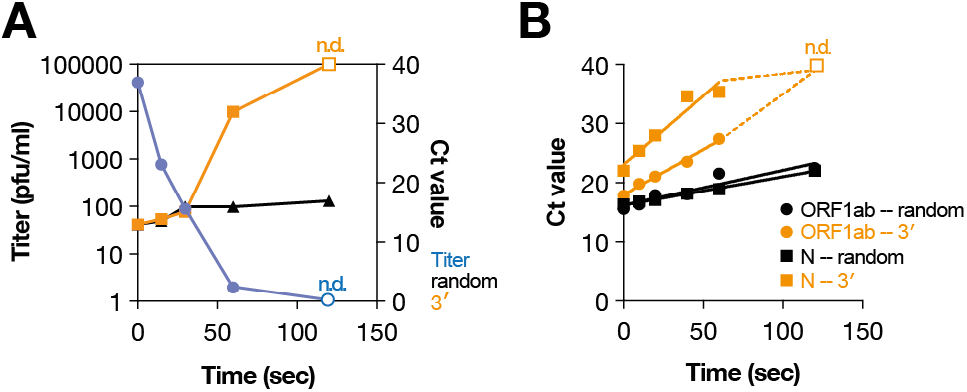
Detection of infectious and non-infectious coronavirus RNA. (**A**) Relation between UVC exposure at 54 W, the HCoV 229E titer, and the qPCR signal following RT reactions with random hexamers or a universal influenza A virus primer capable of binding to the 3′ terminus of each of the eight viral RNA segments. qPCR was performed using primers specific for the NA segment. Plaque assay results that were not detectable (n.d.; open squares) are indicated. RT-qPCR signals that were not detectable are shown as Ct = 40 on graph. (**B**) RT-qPCR of SARS-CoV-2 RNA following UVC exposure for 5, 10, 15, 20, 40 or 60 seconds at 54 W UVC. Data were fit with linear regression. Signals that were not detectable (n.d.) are indicated with open squares.

### SARS-CoV-2 RNA detection in vitro

To investigate if the long-range RT-qPCR protocol can also differentiate between infectious and non-infectious SARS-CoV-2 RNA, SARS-CoV-2 RNA was exposed to 54 W of UVC for 0 to 120 sec. cDNA molecules were generated using random hexamers or an oligo-dT_20_ primer capable of binding to the 3′ polyA-tail of the SARS-CoV-2 RNA genome. Next, we performed qPCR analyses using CDC-recommended primer sets targeting the N and ORF1ab coding regions. For cDNA samples generated using the random hexamers, we observed a relatively limited change in the qPCR Ct value with both primer sets (Fig. 3B). However, a clear change in the qPCR Ct value as function of the exposure time was observed for cDNA samples generated with the oligo-dT_20_ primer for both primer sets (Fig. 3B), indicating that the long-range method can distinguish between intact and infectious SARS-CoV-2 RNA, and fragmented and non-infectious SARS-CoV-2 RNA, in line with the HCoV 229E results (Fig. 1A).

### SARS-CoV-2 RNA detection in clinical samples

To investigate if our long-range RT-qPCR method worked on clinical samples using clinically approved COVID-19 RT-qPCR reagents, we designed a two-step protocol based on the Thermo Fisher TaqPath kit. First, total RNA was extracted from clinical samples using Trizol. Second, cDNA was generated by incubating the extracted RNA with either random hexamer or oligo-dT_20_ and reverse transcriptase. Finally, the cDNA was mixed with the TaqPath master mix, heated to 95 degrees Celsius to denature the cDNA and inactivate the RT in the TaqPath master mix, and analyzed using the TaqPath protocol according to the manufacturer’s instructions. When we used this straightforward protocol to analyze COVID-19 clinical samples (see table S1 for diagnostic values obtained with CoviPath one-step RT-qPCR kit) before UVC exposure, we observed no significant difference between the two methods, indicating that the sensitivity of the long-range RT-qPCR assay was not significantly different from the random hexamer RT-qPCR. Next, we analyzed the samples after UVC exposure and observed a significant difference between the samples analyzed using the long-range RT-qPCR method compared to the samples analyzed using the standard RT-qPCR protocol (Fig. 4A). Because the mean Ct value of the long-range RT-qPCR assay was higher than the mean Ct value of the standard RT-qPCR protocol, these findings indicate that the long-range RT-qPCR assay provides a better estimate of the SARS-CoV-2 RNA integrity than the standard RT-qPCR assay.

**Figure 4.**
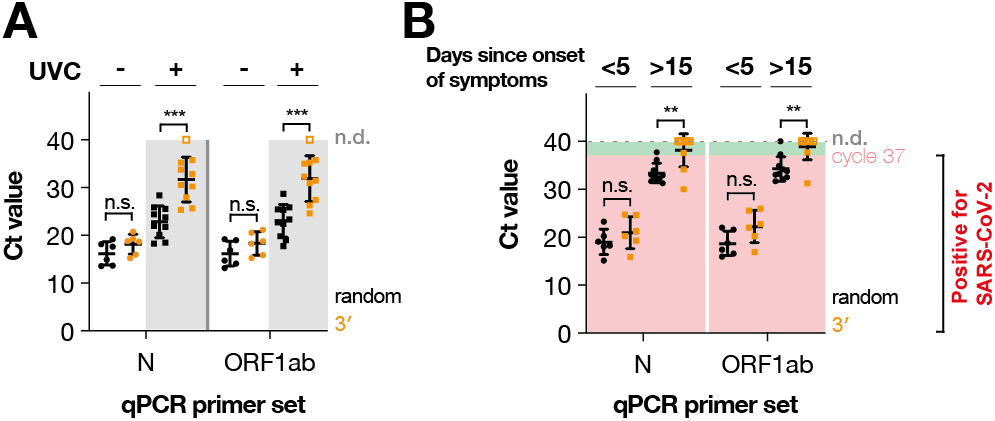
Differential detection of SARS-CoV-2 RNA. (**A**) RT-qPCR of COVID-19 clinical samples following 0 or 120 seconds of 54W of UVC exposure. (**B**) RT-qPCR of COVID-19 clinical samples obtained within 5 days of the onset of clinical symptoms or after 15 days following the onset of clinical symptoms. RT-qPCR signals that were not detectable are shown as open squares at Ct = 40 on graph. Significance was tested using one-way ANOVA with multiple corrections. Not significant (n.s.), p < 0.01 (**), p < 0.001 (***).

### SARS-CoV-2 RNA detection over the course of an infection

To investigate if the two-step, long-range RT-qPCR COVID-19 detection could be used to analyze SARS-CoV-2 RNA levels over the course of a SARS-CoV-2 infection, we compared the above long-range protocol to a clinically approved one-step Taq-Path protocol on clinical samples obtained within the first 5 days of the onset of symptoms (i.e., when SARS-CoV-2 infected patients shed infectious virus). Samples were obtained from COVID-19 patients who were non-immunocompromised. First, RNA was extracted from clinical samples using the MagMAX Viral/Pathogen Nucleic Acid Isolation Kit. Next, either a one-step Taq-Path RT-qPCR protocol was performed or the above two-step RT-qPCR protocol. As shown in Fig. 4B and Table S2, we observed no significant difference between the two RT-qPCR methods for these COVID-19 samples, indicating that the long-range protocol can detect SARS-CoV-2 RNA in clinical samples with a sensitivity that is equal to the one-step protocol, in line with our results in Fig. 4A.

Next, we compared the two-step, long-range RT-qPCR protocol to the one-step Taq-Path protocol on samples obtained at least 15 days after the onset of symptoms when the above patients had recovered. Based on recent human experimental model data, these patients are typically free of infectious virus (6). In line with our in vitro data (Fig. 2 and 3), we observed a significant difference between the two protocols (Fig. 4B). Specifically, we found that 7 out of 10 long-range RT-qPCR reactions produced no detectable signals for both of qPCR primer sets in the TaqPath kit, and only 2 out of the 10 reaction produced signals for both qPCR primer sets. In contrast, all (n=10) RT-qPCR reactions that were performed using the random hexamers in the TaqPath kit returned Ct values below the positivity cut-off (Ct ≤ 37) for at least one of the qPCR primer sets (Table S3).

## Discussion

Current clinical RT-qPCR tests cannot differentiate between RNA from infectious viruses or residual viral genome fragments that are not infectious. In several percent of COVID-19 patients, such residual viral RNAs can be detected long after recovery and the disappearance of infectious SARS-CoV-2 (3–5). These “persistently-positive” RT-qPCR results can lead to incorrect conclusions about the status of a COVID-19 patient. We here used primers that bind to the 3′ terminus of full-length influenza A virus, HCoV 229E, or SARS-CoV-2 genome RNA molecules to reduce the detection of non-infectious viral RNA by RT-qPCR. Our results confirm previous observations with influenza A viruses (7, 8) showing that long-range RT-qPCR data more closely match measurements of infectious virus levels than RT-qPCR data obtained with random hexamers. Moreover, we demonstrate that the long-range RT-qPCR has a similar sensitivity as a commercially available and clinically approved, high-throughput detection assays. Although we use a two-step protocol here, our protocol can easily be converted to a one-step protocol by suppliers of RT-qPCR kits if they replace the random hexamers with an oligo-dT20 primer, ensuring that high-throughput can be achieved. Overall, our data suggest that the long-range RT-qPCR method would be suitable for verifying positive RT-qPCR results, and making a more informed decision about the infectiousness of COVID-19 patients.

## Methods

### Ethics nasopharyngeal swabs

Nasopharyngeal swabs for UVC inactivation study were obtained by Krsnaa Diagnostics Ltd. (KDL). Our use of these swabs was reviewed by the National Accreditation Board for testing and calibration Laboratories (NABL) and the authorized signatories of KDL for the Late. Jayabai Nanasaheb Sutar Hospital, and approved on 22 June 2021. KDL is accredited by the NABL with ISO 15189:2012 compliance and approved by ICMR, with ICMR code KDPLP. Nasopharyngeal swab samples for the time-course analysis of SARS-CoV-2 infections were obtained from Vilnius Santaros Klinikos Biobank (Vilnius, Lithuania). The investigation was approved by Vilnius Regional Bioethics Committee (approval number 2021/5-1342-818).

### Viruses and cells

HEK 293T, Vero-E6 and MDCK cells were originally sourced from ATCC. Influenza A/WSN/33 (H1N1) virus was produced by transfecting a 12-plasmid rescue system into HEK 293T cells (22). After two days, the P0 virus was amplified on MDCK cells in Minimal Essential Medium (MEM; Gibco) containing 0.5% fetal bovine serum (FBS; Sigma) at 37 °C and 5% CO_2_. P1 and P2 viruses were aliquoted and stored at −70 °C. SARS-CoV-2 Bavpat-1 was grown on Vero-E6 cells in Dulbecco’s Minimal Essential Medium (DMEM; Gibco) containing 0.5% FBS at 37 °C and 5% CO_2_. HCoV 229E was sourced from ATCC, AF304460 and grown on Vero-E6 cells. Viral RNA was extracted using Trizol (Invitrogen) as described previously (23).

### UVC chamber and exposure

For UVC exposure, we used a Suraxa® UVC chamber (Pune Instrumentation Private Limited (PIPL)). The unit contained four 254 nm UVC light tubes (Philips) whose output could be adjusted manually and measured using a UV light power meter placed in the center of the unit. The setting used were 11, 32, 43 or 54 W. Samples were placed in a polystyrene well (TRP product number 92012) and positioned, without lid, on the bottom rack in the middle of the UVC chamber. UVC exposures were subsequently performed as indicated in the results section and figures, typically between 0 and 120 seconds. After UVC exposure, virus was eluted using 275 µl PBS/0.05% Tween-80 (24) and transferred to 1.5 ml tubes for plaque assays.

### Plaque assays

Plaque assays were performed as described previously (24). Briefly, samples were serially diluted in MEM containing 0.5% FBS. Two hundred µl of diluted virus was next added to confluent MDCK cells and incubated for 1 hour 37 °C. After virus adsorption to the MDCK cells, the inoculum was removed and replaced with 2 ml MEM/agarose overlay (MEM, 0.5% FBS, 1% agarose). Plaques were grown for 2 days at 37 °C and then fixed with 4% paraformaldehyde in PBS. Plaques were counter-stained with 0.01% crystal violet in water and washed with tap water before analysis.

### RT-qPCR analysis of influenza A virus and HCoV 229E RNA levels

Viral RNA was extracted from 50 µl of elution material using Trizol (Invitrogen) as described previously (23). Extracted RNA was resuspended in 10 µl water. Next, 1 µl of viral RNA was used for reverse transcription with universal influenza 3′ primer TUMI 12G or random hexamers (Thermo Fisher) and SuperScript III (Invitrogen). qPCR was performed using primers specific for the NA segment (25) and Brilliant III Master Mix with high Rox (Agilent) on a Step-One plus qPCR machine. For HCoV 229E, we performed reverse transcription with oligo-dT_20_ (Thermo Fisher) or random hexamers and SuperScript III. qPCR was performed using primers specific for the N gene (21).

### RT-qPCR analysis of UVC exposed SARS-CoV-2 RNA samples

Viral RNA was extracted from 50 µl of elution material using Trizol (Invitrogen) as described previously (23). Extracted RNA was resuspended in 10 µl water. Next, 1 µl of viral RNA was used for reverse transcription with oligo-dT_20_ (Thermo Fisher) or random hexamers and SuperScript III. qPCR was performed using previously described primers specific for the ORF1ab or N coding regions (26). The qPCR was run and analyzed on a QuantStudio 5 or Step-One plus qPCR machine according to the manufacturer’s instructions.

### RT-qPCR analysis of persistently positive time course samples

Viral RNA was extracted using the MagMAX Viral/Pathogen Nucleic Acid Isolation Kit (Thermo Fisher). Next, 4 µl of RNA was mixed with 1 µl of 50 µM oligo-dT_20_ (Thermo Fisher) and 6.4 µl of water. The primer/RNA mix was incubated for 2-3 min at 95 °C and immediately placed on ice afterwards. cDNA produced using Maxima H minus reverse transcriptase (Thermo Fisher) at 50 °C for 1 hour according to the manufacturer’s instructions. qPCR analysis was performed by adding 5 µl of cDNA to a TaqPath qPCR reaction (Thermo Fisher). The qPCR was started with a 2 min 95 °C denaturation step to inactivate the RT enzyme in the TaqPath kit. This was then followed by 40 cycles of 3 sec 95 °C and 30 seconds at 60 °C. The reactions were analyzed on a QuantStudio 5 qPCR machine.

### Statistics

For statistical testing, we used one-way ANOVA and multiple corrections. Statistical testing was performed using GraphPad Prism 9.0.

### Role of the funding sources

The funders of the study had no role in study design, data collection, data analysis, or writing of the manuscript.

## Data Availability

All data produced in the present study are provided as supplemental tables or available upon reasonable request to the authors

## Author contributions

Conceived study: AJWtV, VG. Designed and performed experiments: AJWtV, CR, DJ, IO. Contributed reagents and equipment: PM, KD, VH, TS, VJ, SM, VG. Reviewed and performed data analysis: AJWtV, DJ, CR, IO, VG. Wrote manuscript draft: AJWtV. Contributed to manuscript finalization: AJWtV, DJ, CR, IO, PM, KD, VH, TS, VJ, SM, HJ, VG.

## Data sharing

Ct values of COVID-19 clinical samples are provided in supplemental tables. Reagents and other data are available upon request.

## Declaration of interests

The authors declare no competing interests.

## Acknowledgments

AJWtV is supported by joint Wellcome Trust and Royal Society grant 206579/Z/17/Z. CR is supported by PhD training grant G107570 from Public Health England awarded to AJWtV and HJ.

## Notes

### Competing Interest Statement

The authors have declared no competing interest.

### Funding Statement

This study was funded by joint Wellcome Trust and Royal Society grant 206579/Z/17/Z, and grant G107570 from Public Health England.

### Author Declarations

The Vilnius Regional Bioethics Committee approved this study with samples from the Vilnius Santaros Klinikos Biobank under number 2021/5-1342-818. The National Accreditation Board for testing and calibration Laboratories (NABL) approved this study under code KDPLP on 22 June 2021.

### Summary of Updates

Corrected title, clarified methods, and added new data.

