## Supplemental tables for "Differentiating between infectious and non-infectious influenza A virus and coronavirus RNA levels using long-range RT-qPCR"

**Table S1**. Clinical Ct values of samples used for Fig. 4A.

| | **CoviPath 1-step multiplex kit** | |
|---|---|---|
| **Sample ID** | **Target** | **Ct** |
| 1 | N | 17.7 |
| | ORF1ab | 16.91 |
| 2 | N | 19.27 |
| | ORF1ab | 19.27 |
| 3 | N | 14.97 |
| | ORF1ab | 14.11 |
| 4 | N | 13.74 |
| | ORF1ab | 13.17 |
| 5 | N | 13.55 |
| | ORF1ab | 14.72 |
| 6 | N | 18.16 |
| | ORF1ab | 18.98 |
| 7 | N | 19.01 |
| | ORF1ab | 18.85 |
| 8 | N | 16.25 |
| | ORF1ab | 16.66 |
| 9 | N | 16.39 |
| | ORF1ab | 17.41 |
| 10 | N | 21.14 |
| | ORF1ab | 21.37 |

**Table S2**. Ct values of samples takes <5 days after onset of symptoms. Samples 3-6 were excluded from Fig. 1 and statistical analysis because of mismatched probes.

| | **oligodT$_{20}$ primer** | | **Random hexamers** | |
|---|---|---|---|---|
| **Sample ID** | **Target (probe)** | **Ct** | **Target (probe)** | **Ct** |
| 1 | N (TaqPath) | 19.3 | N (TaqPath) | 15.2 |
| | ORF1ab (TaqPath) | 20.3 | ORF1ab (TaqPath) | 16.0 |
| 2 | N (TaqPath) | 24.7 | N (TaqPath) | 23.3 |
| | ORF1ab (TaqPath) | 26.0 | ORF1ab (TaqPath) | 22.0 |
| 3 | N (TaqPath) | 22.8 | N (LifeRiver) | 20.8 |
| | ORF1ab (TaqPath) | 23.7 | ORF1ab (LifeRiver) | 20.9 |
| 4 | N (TaqPath) | 14.9 | N (GeneXpert) | 14.8 |
| | ORF1ab (TaqPath) | 15.9 | E (GeneXpert) | 12.4 |
| 5 | N (TaqPath) | 25.3 | N (GeneXpert) | 24.5 |
| | ORF1ab (TaqPath) | 25.9 | E (GeneXpert) | 22.6 |
| 6 | N (TaqPath) | 24.6 | N (GeneXpert) | 23.9 |
| | ORF1ab (TaqPath) | 25.9 | E (GeneXpert) | 22.1 |
| 7 | N (TaqPath) | 20.3 | N (TaqPath) | 18.3 |
| | ORF1ab (TaqPath) | 22.0 | ORF1ab (TaqPath) | 17.4 |
| 8 | N (TaqPath) | 21.3 | N (TaqPath) | 20.3 |
| | ORF1ab (TaqPath) | 22.8 | ORF1ab (TaqPath) | 21.6 |
| 9 | N (TaqPath) | 15.9 | N (TaqPath) | 18.1 |
| | ORF1ab (TaqPath) | 16.9 | ORF1ab (TaqPath) | 16.6 |
| 10 | N (TaqPath) | 24.6 | N (TaqPath) | 19.1 |
| | ORF1ab (TaqPath) | 25.6 | ORF1ab (TaqPath) | 18.8 |

**Table S3**. Ct values of samples takes >15 days after onset of symptoms. Undetectable levels are indicated with U.D.

| | oligod$T_{20}$ primer | | Random hexamers | |
|---|---|---|---|---|
| **Sample ID** | **Target (probe)** | **Ct** | **Target (probe)** | **Ct** |
| 1 | N (TaqPath) | UD | N (TaqPath) | 33.0 |
| | ORF1ab (TaqPath) | UD | ORF1ab (TaqPath) | 32.5 |
| 2 | N (TaqPath) | UD | N (TaqPath) | 32.8 |
| | ORF1ab (TaqPath) | UD | ORF1ab (TaqPath) | 33.8 |
| 3 | N (TaqPath) | 37.7 | N (TaqPath) | 31.7 |
| | ORF1ab (TaqPath) | UD | ORF1ab (TaqPath) | 33.3 |
| 4 | N (TaqPath) | 30.0 | N (TaqPath) | 32.1 |
| | ORF1ab (TaqPath) | 31.3 | ORF1ab (TaqPath) | 33.1 |
| 5 | N (TaqPath) | 34.2 | N (TaqPath) | 32.3 |
| | ORF1ab (TaqPath) | 37.8 | ORF1ab (TaqPath) | 32.1 |
| 6 | N (TaqPath) | UD | N (TaqPath) | 31.3 |
| | ORF1ab (TaqPath) | UD | ORF1ab (TaqPath) | 31.7 |
| 7 | N (TaqPath) | UD | N (TaqPath) | 37.8 |
| | ORF1ab (TaqPath) | UD | ORF1ab (TaqPath) | 37.3 |
| 8 | N (TaqPath) | UD | N (TaqPath) | 36.2 |
| | ORF1ab (TaqPath) | UD | ORF1ab (TaqPath) | 34.9 |
| 9 | N (TaqPath) | UD | N (TaqPath) | 34.0 |
| | ORF1ab (TaqPath) | UD | ORF1ab (TaqPath) | UD |
| 10 | N (TaqPath) | UD | N (TaqPath) | 33.1 |
| | ORF1ab (TaqPath) | UD | ORF1ab (TaqPath) | 34.2 |
